# Short of Breath for the Long Haul: Diaphragm Muscle Dysfunction in Survivors of Severe COVID-19 as Determined by Neuromuscular Ultrasound

**DOI:** 10.1101/2020.12.10.20244509

**Authors:** Ellen Farr, Alexis R. Wolfe, Swati Deshmukh, Leslie Rydberg, Rachna Soriano, James M. Walter, Andrea J. Boon, Lisa F. Wolfe, Colin K. Franz

## Abstract

Many survivors from severe coronavirus disease 2019 (COVID-19) suffer from persistent dyspnea and fatigue long after resolution of the active infection. In a cohort of 25 consecutive COVID-19 survivors admitted to an inpatient rehabilitation hospital (76% male), 80% of them had at least one sonographic abnormality of diaphragm muscle structure or function.

Specifically, when compared to established normative data, 76% had reduced diaphragm thickening ratio (impaired contractility), and 20% patients had reduced diaphragm muscle thickness (atrophy). These findings support neuromuscular respiratory dysfunction as a highly prevalent underlying cause for prolonged functional impairments after hospitalization for COVID-19.

## Introduction

The substantial burden of chronic disability post-hospitalization for coronavirus disease 2019 (COVID-19) is increasingly clear.^1^ In a recent study, nearly half of COVID-19 patients were not able to return to work 60 days after hospital discharge.^1^ Survivors of severe COVID-19 frequently report persistent shortness of breath, cough and fatigue post-hospitalization.^1, 2^ While these symptoms may stem from direct involvement of the lung parenchyma itself, the possibility of underlying neuromuscular respiratory weakness should be considered. Neurological manifestations of COVID-19 are increasingly recognized with prominent involvement of the neuromuscular system ranging from mild creatine kinase (CK) elevation to flaccid tetraplegia requiring tracheostomy.^3^ Here we report neuromuscular ultrasound findings that define the unexpectedly high prevalence of structural and functional alterations to the diaphragm muscle after hospitalization for COVID-19.

## Methods

Consecutive patients (n=25) were identified on admission to a dedicated COVID-19 unit at a single rehabilitation hospital (Shirley Ryan AbilityLab, Chicago, USA). They were admitted to inpatient rehabilitation from 14 separate acute care hospitals between July 21^st^, 2020 and September 24^th^, 2020. Acute care data was obtained as available through chart review of medical records as summarized in table 1. The diaphragm muscle was assessed on a portable ultrasound system (TE7, Mindray, Mahwah, USA) with either a 6-to 14-MHz linear array or 2-to 5-MHz curvilinear array selected to maximize image clarity on the basis of individual characteristics such as body habitus. Briefly, each hemi-diaphragm was identified in the zone of apposition, and thickness was measured at end-expiration and maximal inspiration (figure 1). Normal values have been established for diaphragm end-expiratory muscle thickness (>0.14 cm) and thickening ratio (>1.2), calculated as thickness at maximal inspiration/thickness at end-expiration.^4^ Approval obtained from Northwestern University IRB (STU23625789).

**Table 1.** Summary of key patient characteristics.

| **Table 1** Baseline characteristics, lab values, and lengths of stay of post-COVID-19 patients in inpatient rehabilitation |  |
| --- | --- |
| **Subject Characteristics** |  |
| **Age** | 59.1±13.2 (33-89) |
| **Sex M:F (n=25)** | 19:6 |
| **BMI** | 30.8±6.1 (21-48) |
| **Comorbidities** | Hypertension 68% (17/25)<br>Diabetes Mellitus or Pre-Diabetes Mellitus 56% (14/25)<br>Hyperlipidemia 30.4% (7/25)<br>Asthma and/or COPD 20% (5/25)<br>Cancer 13.4% (3/25)<br>Cerebrovascular Accident 13.4% (3/25)<br>Acquired Immunodeficiency 13.4% (3/25) |
| **Days in ICU** | 38±21.7 (2-83) |
| **Days Hospitalized** | 45.6±25.5 (10-92) |
| **Days of Mechanical Ventilation** | 43.1±33.5 (10-153) |
| **Days Since Onset\*** | 72.2±46.5 (3-184) |
| **Days Since ICU** | 32.4±22.9 (2-80) |
| **CRP (mg/L)** | 180.2±135.9 (3.5-423) |
| **Troponin (ng/mL)** | 0.7±2.0 (0.003-8.09) |
| **CK (U/L)** | 753.3±976.2 (39-3090) |
| **A1c (%)** | 7.9±2.5 (5.1-13.7) |

Mean ± SD (Range) or % as appropriate
\*As defined by first positive test, onset of symptoms, or hospital admission where available

**Figure 1.**
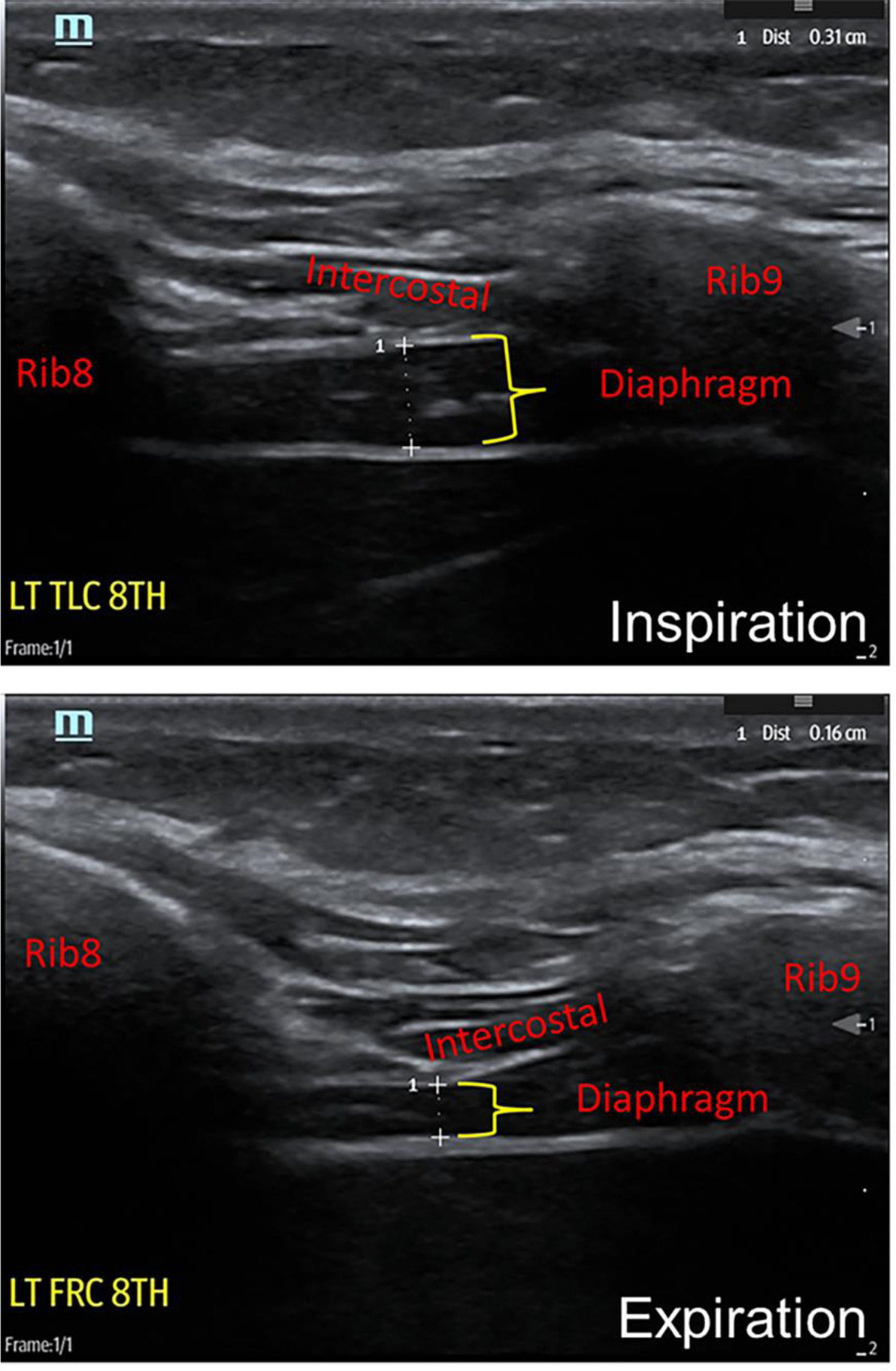
Technique for neuromuscular ultrasound examination of hemi-diaphragm. Top panel, thickness of diaphragm at end inspiration or total lung capacity (0.31 cm). Bottom panel, thickness of diaphragm at maximal expiration or functional residual capacity (0.16cm). The thickening ratio was calculated at 1.94 (0.31cm/0.16cm).

## Results

This report summarizes the assessment of 25 consecutive patients who survived COVID-19 and were admitted to an inpatient rehabilitation unit in table 1. Briefly, of these patients 6 (24%) were female and 19 (76%) were male, 12 (48%) had a diagnosis of pre-diabetes or diabetes mellitus, 15 (60%) had hypertension, whereas only 3 (12%) had a diagnosis of chronic obstructive pulmonary disease (COPD) or asthma. The average time spent on a ventilator at the acute care hospital was 44.3 days and the mean peak CK level in these patients was 753 U/L (range 39-3090 U/L). A total of 10 patients (40%) were on supplemental oxygen at the time of diaphragm ultrasound, and none of them were ventilator dependent. There were 5 patients (20%) who had an end-expiration thickness value below the normal cut off, whereas 19 patients (76%) had a reduced thickening ratio on at least one side, and 10 patients (49%) had reduced thickening ratio bilaterally. Overall, 20 patients (80%) had at least one structural or functional abnormality on diaphragm ultrasound (figure 2).

**Figure 2.**
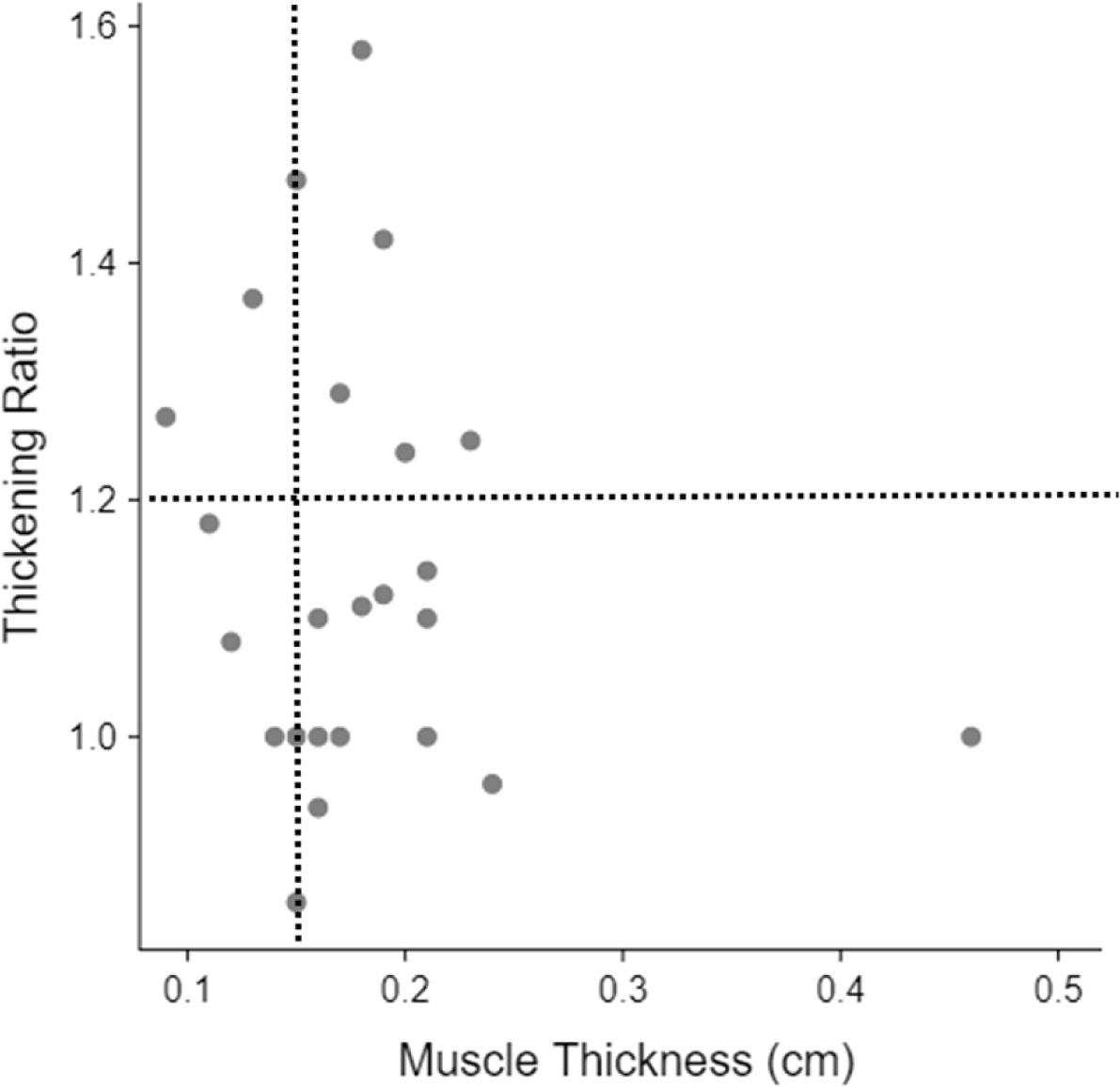
Sonographic diaphragm muscle thickness versus thickening ratio in survivors of COVID-19 that require inpatient rehabilitation. Individual cases plotted to the left of the vertical dotted line have abnormal thickness (i.e. muscle atrophy). Those plotted below the horizontal dotted line have abnormal thickening ratio (i.e. impaired contractility). Taken together, only the 5 cases (out of 25) located in the upper right quadrant fall within normal limits across both parameters.

## Discussion

To our knowledge this is the first study utilizing ultrasound to evaluate the diaphragm in COVID-19 survivors and demonstrates that patients who require inpatient rehabilitation after acute hospitalization have a very high prevalence of diaphragm dysfunction. These findings are remarkably similar to a previous report of sonographically identified diaphragm dysfunction in patients with confirmed myopathy.^5^ In contrast, diaphragm ultrasound findings in patients with chronic dyspnea related to COPD largely resembled the healthy population.^6^ Given the large number of COVID-19 survivors who suffer from persistent dyspnea months after onset of disease,^7^ it is possible that diaphragm muscle dysfunction is a major contributing factor.

Recent autopsy data obtained from COVID-19 patients admitted to an intensive care unit in the Netherlands demonstrated that the angiotensin-converting enzyme 2 (ACE-2) receptor represents an entry point for the severe acute respiratory syndrome coronavirus 2 to directly infect the diaphragm.^8^ However, only a modest subset (4 out of 26) had detectable levels of viral ribonucleic acid present in the muscle tissue. Therefore, we hypothesize that the mechanism of diaphragm involvement for the majority of COVID-19 patients is not from direct muscle infection, but more likely analogous to that of critical illness myopathy (CIM). The pathophysiology of CIM is considered a multifactorial condition associated with exposure to high-dose steroids, muscle membrane dysfunction, microcirculatory changes associated with inflammation, and impaired glucose transporter type 4 translocation to the muscle membranes.^9, 10^ This combination of factors results primarily in atrophy of type 2 muscle fibers. The diaphragm is likely injured via the same mechanism in COVID-19 as it is comprised of a nearly even mix of type 1 and type 2 myofibers.^11^

Our study provides new insight into neuromuscular respiratory weakness as an important contributor to prolonged functional impairments in survivors from COVID-19. It remains unclear to what extent diaphragm involvement may exist in milder COVID-19 not requiring hospitalization. Subjective dyspnea and fatigue are among the most frequently reported symptoms in these so called “long haulers”.^12^ Clinicians should consider a diaphragm ultrasound study if there is clinical concern for neuromuscular respiratory weakness in non-hospitalized patients with COVID-19. Currently, long term dysfunction in these patients is primarily attributed to lung parenchymal damage, overlooking diaphragm dysfunction as a possible contributor.^13^ Given that the diaphragm is the main respiratory muscle, we speculate that prolonged dysfunction may be a factor in at least a subset of patients. The high prevalence of diaphragm dysfunction in more severe COVID-19 survivors highlights the importance of neuromuscular respiratory rehabilitation protocols in this population. Further investigation is required to determine how diaphragm structure and function recover after hospital discharge and how this may related to their functional status or risk for further medical complications.

## Data Availability

The de-identified clinical data presented here will be made available to academic-based medical and health researchers on request.

## Abbreviations list

(COVID-19): Coronavirus disease 2019
(CK): Creatine Kinase
(COPD): Chronic Obstructive Pulmonary Disease
(CIM): Critical Illness Myopathy
(ACE-2): Angiotensin-Converting Enzyme 2

